# Impaired Adaptive Learning in Chronic Pain Contributes to Apathy

**DOI:** 10.1101/2025.10.20.25336624

**Authors:** Xinyuan Yan, Crina M. Peterson, Lisa M. Schmidt, Seth Koenig, Donald R. Nixdorf, Alexander Herman, David P. Darrow

**Affiliations:** Department of Neurosurgery, University of Minnesota, Minneapolis, MN, United States; Department of Psychiatry, University of Minnesota, Minneapolis, MN, United States; The TMJ Association, Milwaukee, WI, United States; School of Dentistry, University of Minnesota, Minneapolis, MN, United States

## Abstract

The cognitive mechanisms linking chronic pain to motivational symptoms remain poorly understood. We demonstrate that individuals with chronic temporomandibular disorder (TMD), a common cause of chronic pain, exhibit a specific deficit in adaptive learning in uncertain environments, characterized by failure to reduce uncertainty over time and maintain efficient learning rates. Using a probabilistic reward task, we pioneered the application of a novel volatile Kalman filter to model behavior in 26 TMD participants and 39 matched controls, uniquely tracking trial-wise updates in uncertainty, volatility, and learning rate. Although surface-level performance did not differ across groups, model-based analysis revealed that those with TMD failed to reduce uncertainty and adapt their learning over time. TMD participants also reported significantly greater apathy, depression, and pain catastrophizing, as well as lower health-related quality of life. Mediation analysis confirmed that impaired uncertainty adaptation partially mediated the relationship specifically between TMD and apathy. These findings identify a computational signature of disrupted uncertainty adaptation in people with TMD and provide evidence for a mechanistic link between chronic pain and motivational dysfunction. This work lays a foundation for future studies examining how belief-updating deficits contribute to broader affective and cognitive symptoms in chronic pain.

## Introduction

Chronic pain affects 21% of the population and causes substantial morbidity and disability. Although often treated as peripheral in origin, a growing body of evidence suggests that chronic pain involves central mechanisms linked to emotion, cognition, and motivation (Apkarian et al., 2009; Baliki and Apkarian, 2015). In particular, perceptions of uncertainty may play an important role in the burden experienced by people with chronic pain (Brown et al., 2008; Wiech and Tracey, 2009; Yoshida et al., 2013). Uncertainty about diagnosis, prognosis, and treatment efficacy can amplify pain perception, increase anxiety, and lead to maladaptive coping mechanisms. For many patients, not knowing what causes their symptoms or when pain might intensify creates a state of hypervigilance that can exacerbate muscle tension and pain. This uncertainty often leads to catastrophizing thoughts, where patients anticipate the worst possible outcomes, creating a self-perpetuating cycle of stress and pain that can leave patients feeling helpless and apathetic (Quartana et al., 2009; Vlaeyen and Linton, 2000). Clinicians working with people who have chronic pain have long known that addressing altered perceptions of these uncertainties may help reduce anxiety and improve outcomes.

One particularly disabling form of chronic pain, temporomandibular disorders (TMD), affects 5% to 10% of the population and is the second most common cause of chronic musculoskeletal pain(National Academies of Sciences, Engineering, and Medicine et al., 2020). Anxiety and depression, conditions associated with low motivation, are found in 20-60% of patients with TMD (De La Torre Canales et al., 2018), similar to the prevalence rates seen in chronic pain more generally (Aaron et al., 2025). Patients with TMD exhibit executive function deficits related to the functional disconnection of the cognitive, motivational, and affective areas of the brain (Weissman-Fogel et al., 2011). Whether these deficits reflect generalized impairment or disruption in specific cognitive processes that represent potential therapeutic targets remains unknown despite the use of cognitive behavioral therapy as a standard treatment (Shivakumar et al., 2025). Previously, we found that perceptions of uncertainty influence motivation levels in a large normative sample (Yan et al., 2025). Here we provide behavioral and computational evidence that individuals with TMD exhibit a core deficit in adaptive learning in response to uncertainty, which contributes to motivational dysfunction, particularly apathy.

## Results

We recruited 26 individuals with chronic TMD (age±SD=33.69±14.45) and 40 healthy controls (age±SD = 32.60±13.68). One control participant reported unexpectedly high pain (VAS=68/100) despite no diagnosed pain conditions and was excluded as an outlier, yielding a final control sample of 39 participants (age±SD=32.75±13.81). After exclusion, control participants showed minimal pain levels (VAS=6.44±9.76) compared to TMD participants (VAS=36.58±20.64, t(63)=7.91, p<0.001). All TMD participants had a muscle pain diagnosis (9 local myalgia, 17 myofascial pain with referral), and 96% had a joint pain diagnosis (7 unilateral, 18 bilateral arthralgia). The distribution of Graded Chronic Pain Scale (GCPS) was I=11 (42%), II= (27%), III=6 (23%), and IV=2 (8%), which is similar to the distribution in U.S. dental practice settings(Velly et al., 2022) and is composed of primarily mild to moderate severity. Participants completed a 300-trial three-armed bandit task (Figure 1, Method) designed to probe learning under uncertainty. Participants with TMD reported higher pain at baseline (t(63)=7.91, p<0.001) (Figure 1D) and completed self-report assessments of apathy, mood, and pain catastrophizing (details see Method). Task behavior was analyzed using a volatile Kalman filter model (Figure 1C, Method), which estimates latent learning parameters on a trial-by-trial basis.

**Figure 1.**
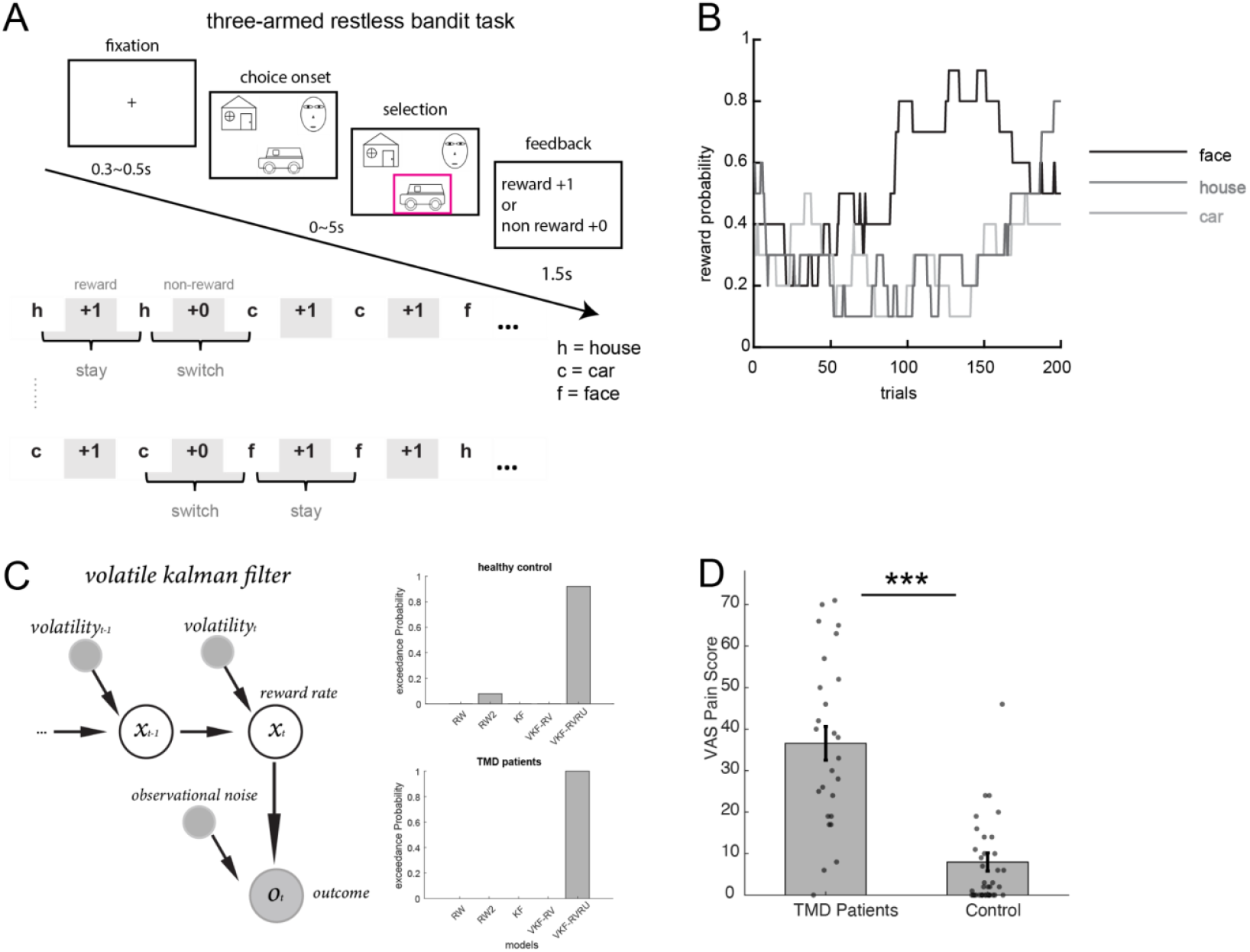
Task, computational models, and pain ratings. (A) Three-armed restless bandit task (Method). Patients chose one of three options (face, car, house), followed by reward or no-reward feedback. (B) Example reward probability trajectories across trials. (C) Volatile Kalman filter model implementation (left panel) and Bayesian model comparison results (right two panels). The model computes trial-by-trial value and uncertainty estimates for each bandit. The volatile Kalman filter with relative value and relative uncertainty incorporated into the softmax function outperformed other models. Model abbreviations: RW = Rescorla-Wagner; Dual RW = Rescorla-Wagner with positive and negative learning rates; KF = Kalman filter; VKF-RV+RU = volatile Kalman filter with relative value and uncertainty in softmax function; VKF-RV = volatile Kalman filter with relative value in softmax function (model validation analyses see Method). Relative value (RV) and relative uncertainty (RU) were calculated as the ratio of the chosen option’s value/uncertainty to the sum across all options (see Method). (D) TMD patients show significantly higher pain ratings on the visual analog scale (VAS) than controls. ***p<0.001.

### TMD participants failed to adapt to uncertainty

Although participants with TMD performed similarly (all p > 0.400, see Method) to healthy controls in model-free measures (e.g., accuracy, performance, win-stay/lose-shift), model-based analyses revealed persistent deficits in learning (Figure 2). Compared to controls (HC), participants with TMD failed to reduce their uncertainty estimates and adapt their learning rates over time (Figure 2A–C). Regression analysis showed that controls decreased their learning rates over time, with significant negative slopes in uncertainty and volatility estimates (all p<0.001), while TMD participants exhibited flat or non-significant trajectories (Figure 2D–F). This pattern suggests TMD participants maintain a chronically heightened sensitivity to environmental changes. First, they fail to decrease their estimate of environmental volatility despite substantial experience. Second, they maintain high uncertainty about reward expectations throughout the experiment. Third, their persistently elevated learning rates indicate that they continually prioritize recent feedback over prior beliefs. This computational signature resembles a trauma-like state where the individual anticipates ongoing volatility and remains hypervigilant to environmental changes. Such a persistent state of uncertainty about the environment may undermine the ability to form stable expectations, contributing to learned helplessness and apathy observed in chronic pain conditions (Samwel et al., 2006).

**Figure 2.**
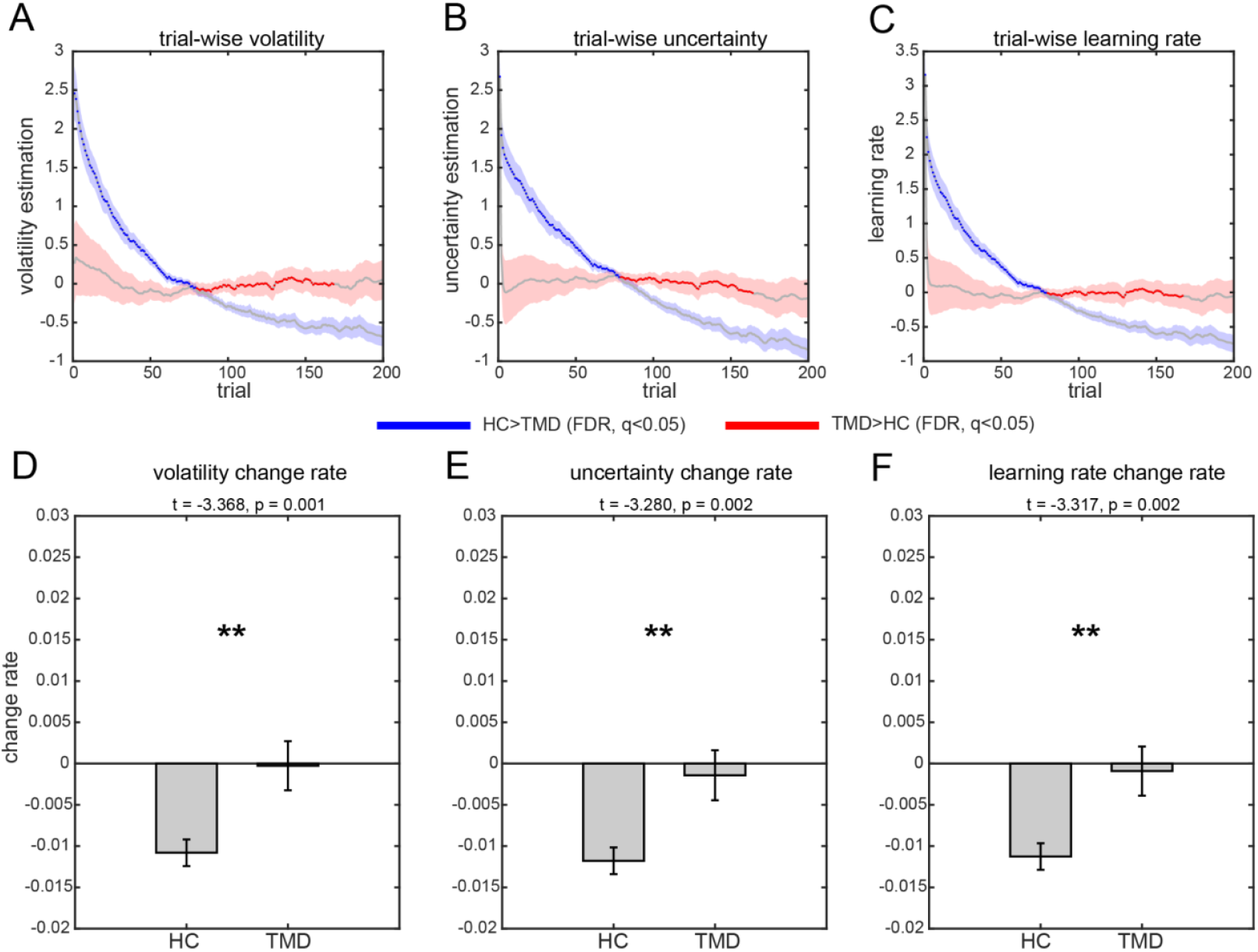
Temporal dynamics of model parameters. (A–C) Trial-by-trial estimates of volatility, uncertainty, and learning rate in TMD and control groups. (D–F) Linear regression slopes for each parameter over time. Controls showed adaptive decreases in parameter values (all p<0.001); TMD participants did not (all p>0.05). Controls also showed significantly decreasing trends than TMD patients as well (all p < 0.01). **p<0.01, ***p<0.001. n.s, non-significant.

To ensure the robustness of these findings, we also tested exponential decay models (see Methods & Table S2 for details). This analysis revealed that TMD participants not only failed to show expected exponential decay but often exhibited paradoxical patterns. For volatility, 82% of controls showed adaptive exponential decay versus only 42% of TMD participants (Fisher’s exact p = 0.001), with 39% of TMD participants showing exponential growth. The decay rate parameter for uncertainty differed significantly between groups (HC: b = -0.00094 vs TMD: b = +0.00012, two-sample t test, p = 0.002), with TMD’s positive value indicating increasing uncertainty over time. For learning rate, 69% of controls versus 35% of TMD participants showed adaptive decay (Fisher’s exact p = 0.01). These converging analyses, whether using linear slopes or exponential parameters, demonstrate that TMD participants exhibit qualitatively aberrant learning trajectories incompatible with adaptive uncertainty processing.

Together, this computational profile, where uncertainty remained unchanged rather than resolved with experience, aligns with predictive-processing (Bayesian) accounts of pain that emphasize aberrant precision (uncertainty) weighting and misestimation of environmental volatility, and with evidence that people learn the statistics of pain in Bayesian-like ways (Onysk et al., 2024). It is also consistent with patients’ lived reports that chronic pain is increasingly unpredictable, which fosters ongoing uncertainty in day-to-day functioning (Little et al., 2025).

### Uncertainty learning dynamics predict psychological outcomes in TMD participants

To assess the clinical relevance of these findings, we examined the psychiatric symptom burden across groups. TMD participants showed significantly higher apathy (Apathy Motivation Index, t(63)=-3.63, p<0.001) (Ang et al., 2017), depression (PHQ-9, t(63)=-6,63, p<0.001), pain catastrophizing scores (t(63)=-4.15, p<0.001), and lower quality of life (EQ-5D; t(63)=-3.70, p<0.001, Figure 3A–D). Mediation analysis demonstrated that reduced uncertainty adaptation significantly mediated the relationship between TMD status and apathy indirect effect (ab), effect = 0.175, 95%CI = [0.006, 0.484], 5000 sample bootstrap) (Figure 3E), suggesting that disrupted uncertainty learning may underlie motivational symptoms in chronic TMD pain. We did not find any other significant mediation results for PHQ-9 (95%CI = [-0.153, 0.283]), Pain Catastrophizing score (95%CI = [-0.123, 0.395]), or EQ-5D (95%CI = [-0.003, 0.447]).

**Figure 3.**
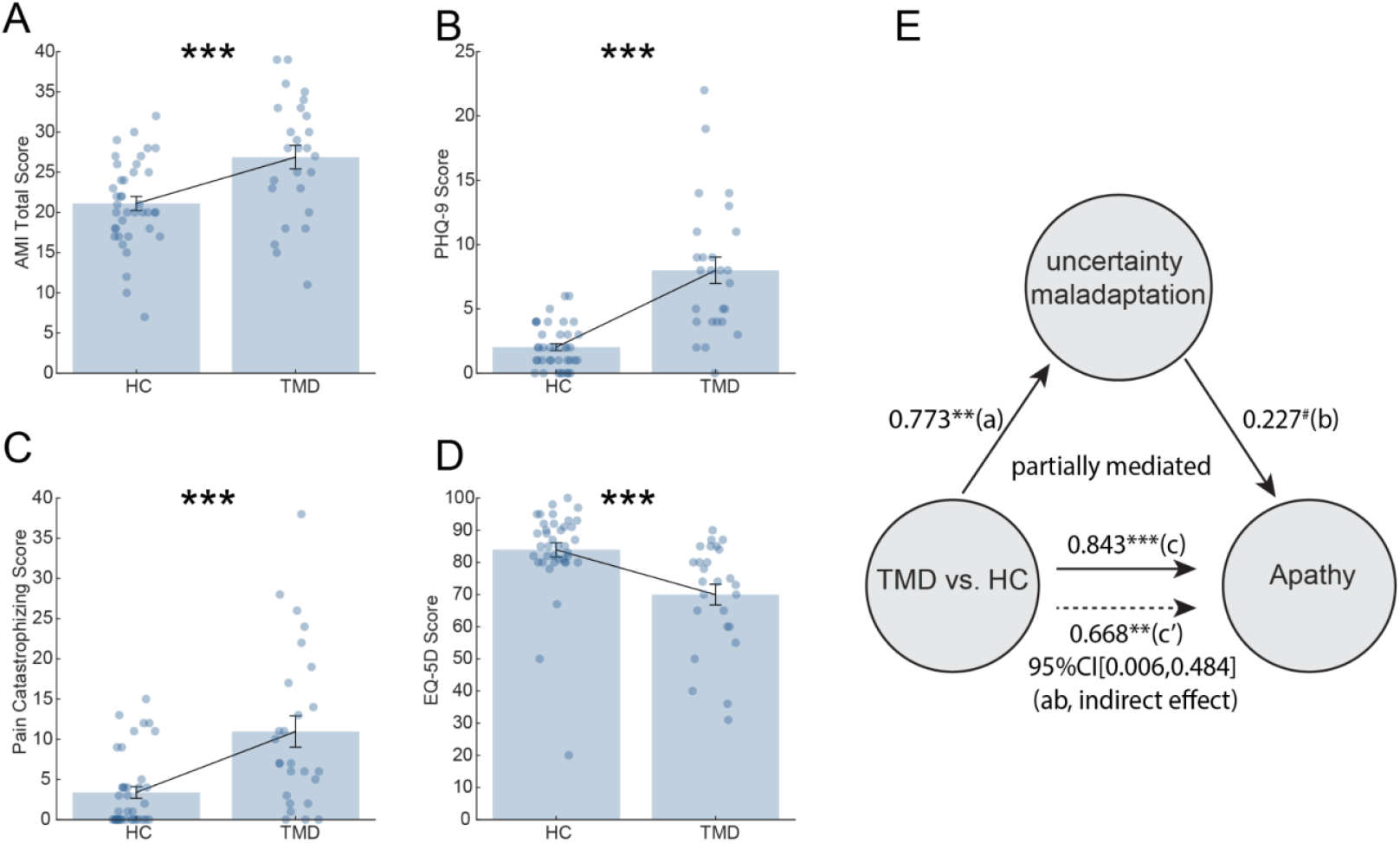
Psychiatric symptom burden and mediation analysis. (A–D) Group differences in apathy, depression, pain catastrophizing, and EQ-5D. (E) The mediation model shows that impaired uncertainty adaptation mediates the relationship between TMD status and apathy (the indirect effect was significant. p<0.06 (marginally significant), *p < 0.05, **p<0.01, ***p < 0.001.

The selective mediation of apathy, but not depression or pain catastrophizing, by uncertainty adaptation deficits reveals a specific computational pathway linking chronic pain to motivational dysfunction. This persistent uncertainty state mirrors the clinical phenomenology of chronic pain, where patients cannot predict when symptoms will worsen, which may lead to learned helplessness and withdrawal.

## Discussion

Our findings identify a computational mechanism that links chronic pain to apathy: impaired adaptation of uncertainty over time. Healthy controls reduced volatility and uncertainty with experience, showing robust exponential decay, whereas participants with temporomandibular disorder (TMD) often failed to reduce uncertainty, and nearly 40% showed paradoxical increases in volatility or uncertainty. This qualitative breakdown suggests that the pain state disrupts belief updating itself, not merely the pace of learning. Such results align with predictive-processing accounts of pain, which propose that maladaptive precision-weighting of sensory and contextual signals sustains hypervigilance and unpredictability(Apkarian et al., 2009; Wiech and Tracey, 2009). Clinically, patients often describe their pain as arising “out of nowhere,” and our findings provide a computational basis for this lived experience.

Mediation analysis further underscored the specificity of this mechanism. Impaired uncertainty adaptation predicted apathy, but not depression, catastrophizing, or quality of life. Apathy is frequently conflated with depression, yet evidence points to distinct neural substrates(Ang et al., 2017; Husain and Roiser, 2018). Our results extend this dissociation into chronic pain, highlighting apathy as the behavioral outcome most directly tied to uncertainty misestimation and suggesting it may serve as a mechanistic bridge between disrupted learning and functional disability.

At the same time, surface performance was preserved. TMD participants tracked rewards above chance, exploited optimal options, and showed normal win–stay/lose–shift behavior. Their impairment emerged only at the computational level, where latent learning parameters failed to adapt. This dissociation explains how patients can appear cognitively intact while struggling with motivation. Together, these findings reframe motivational symptoms in chronic pain as arising from domain-general failures of belief updating rather than nociception or mood symptoms alone. They also suggest a testable therapeutic direction: interventions that recalibrate uncertainty, through cognitive-behavioral strategies or neuromodulation of prefrontal–striatal circuits, may help restore adaptive learning and motivation in patients with chronic pain.

## Materials and Methods

### Three-armed restless bandit task

In this three-armed restless bandit task (Ebitz et al., 2018; Yan et al., 2025), participants completed 200 trials where they freely selected between three targets on each trial to earn potential rewards of 1 point. Each target is associated with a hidden reward probability that randomly and independently changes throughout the task. We seeded each participant’s reward probability randomly to prevent biases due to particular kinds of environments. Specifically, on each correct trial, there was a 10% chance that the reward probability for each target would either increase or decrease by 1.0, with these probabilities bounded between 0.1 and 0.9. Due to the variable and independent nature of the rewards, participants could only estimate the probabilities by actively sampling from the targets and accumulating their reward experiences over time. Trials were excluded if participants failed to respond within 5 seconds. The task was programmed in MATLAB using Psychtoolbox 3 (http://psychtoolbox.org/). Stimuli were presented on an HP laptop positioned approximately 70 cm from the participant’s face and centered in their field of view while they sat upright in their hospital bed.

### Self-reported measures

Participants completed a comprehensive battery of validated self-report measures. The Graded Chronic Pain Scale (GCPS) (Korff et al., 1992) assessed pain intensity and pain-related disability. The Patient Health Questionnaire-9 (PHQ-9) (Kroenke et al., 2001) was administered to evaluate the presence and severity of depressive symptoms. The EuroQol Five Dimension (EQ-5D)(Brooks et al., 2020) questionnaire assessed health-related quality of life, with a higher score indicating better health status. Motivational deficits were evaluated using the Apathy Motivation Index (AMI) (Ang et al., 2017).

### Model-free analyses

We adopted some widely used model-free measures, including win-stay and lose-switch (den Ouden et al., 2013), as the direct measurement for this learning task.

*Win-stay*. Win-stay is the percentage of times that the choice in trial t-1 was repeated on trial t following a reward.

*Lose-switch*. In contrast, the lose-switch equals the percentage of trials in which the choice was shifted or changed when the outcome of trial t-1 was non-reward.

Initial analysis of model-free behavioral metrics, including stay rate, switch rate, win-stay/lose-switch behaviors, overall performance (corrected for chance level), and response times, revealed no significant differences between TMD patients and controls (all p > 0.400; see Figure S1). Given the non-stationary nature of our restless bandit task, we assessed learning through maintenance of above-chance performance and adaptive strategy (see below) use rather than traditional improvement curves, as reward probabilities changed continuously throughout the experiment.

To comprehensively characterize task behavior, we computed seven model-free metrics examining reward sensitivity, choice consistency, and performance optimization (see below).

We first examined choice autocorrelation to assess systematic decision patterns independent of reward feedback. The lag-1 autocorrelation coefficient revealed positive values for both groups (HC: 0.270 ± 0.051; TMD: 0.169 ± 0.064; t(63) = 1.24, p = 0.221), calculated as r = Σ[(choice_t - mean)×(choice_{t+1} - mean)] /Σ(choice - mean)^2^. These positive autocorrelations indicate that both groups exhibited systematic choice patterns rather than random responding, with TMD participants showing a non-significant trend toward lower perseveration that did not reach the statistical threshold.

To assess reward-driven learning, we evaluated how well participants tracked and exploited the objectively best option. Both groups performed well above the chance level of 33.3%, with TMD participants (52.1 ± 3.4%) tracking the best option as effectively as controls (50.4 ± 1.9%; t(63) = -0.49, p = 0.624). Furthermore, we quantified reward-seeking behavior by correlating each participant’s choice probability with each option’s average reward rate. Both groups showed strong positive correlations (HC: 0.522 ± 0.101; TMD: 0.643 ± 0.089; t(63) = -0.84, p = 0.405), with TMD participants displaying numerically higher reward sensitivity, directly contradicting any hypothesis of disengagement or reward insensitivity.

To assess learning success, we calculated individualized chance levels for each participant based on the mean reward probabilities of their specific bandit instantiation. Binomial tests revealed that 66.7% of controls and 61.5% of TMD participants performed above their personalized baseline, with group-level analyses confirming both groups performed significantly above chance (both p < 0.0001). This comparable achievement rate demonstrates that TMD participants successfully learned the task structure and exploited reward opportunities as effectively as controls.

Across all behavioral metrics spanning reward sensitivity, choice consistency, and performance optimization, TMD participants showed normal task behavior with no significant group differences (all p > 0.14). This behavioral equivalence, including reward tracking, systematic choice patterns, and above-chance performance, makes our computational finding particularly specific and interpretable. Despite demonstrating intact trial-by-trial learning and reward processing at the behavioral level, TMD participants uniquely fail to reduce their uncertainty estimates over time, revealing a selective deficit in uncertainty resolution rather than a general learning impairment. This dissociation between preserved behavioral performance and impaired uncertainty updating highlights a subtle but critical difference in how TMD patients process and integrate information under uncertainty.

### Temporal dynamics of model-free behavioral metrics

To examine learning dynamics across the task, we divided each participant’s 200 trials into consecutive 10-trial bins (20 bins total). Within each bin, we calculated win-stay rate (proportion of rewarded trials followed by staying with the same choice) and lose-shift rate (proportion of non-rewarded trials followed by switching to a different choice). These metrics were averaged across participants within each group to generate temporal trajectories. Group differences at each time point were assessed using independent samples t-tests. To examine learning phases, we aggregated data into early (trials 1-50), middle (trials 51-100), and late (trials 151-200) learning periods.

Analysis of behavioral dynamics across 10-trial bins revealed remarkably stable performance in both groups throughout the task (Figure S2). Win-stay rates remained consistently high across all learning phases (early: Controls = 64.0 ± 6.1%, TMD = 67.1 ± 4.2%; late: Controls = 72.6 ± 1.3%, TMD = 71.5 ± 3.8%), with no significant group differences at any time point (0/20 bins, all p > 0.05). Similarly, lose-shift rates showed no group differences across bins (0/20 bins, all p > 0.05), though both groups showed increasing lose-shift rates from early (Controls = 67.8 ± 7.6%, TMD = 71.7 ± 7.0%) to middle learning (Controls = 75.7 ± 5.2%, TMD = 65.3 ± 3.9%).

### Computational modeling

Of note, our approach here is essentially the same as that taken by Piray and Daw ((Chakroun et al., 2020; Piray and Daw, 2020). Here, we briefly described the model details as follows.

#### Kalman filter

The Kalman filter (KF) model has been widely applied in psychology and neuroscience to study various aspects of learning and decision-making (Cheng et al., 2022; Dayan et al., 2000).

In the Kalman filter model for a multi-armed bandit task, *process noise*, and *observation noise* refer to two distinct sources of uncertainty that affect the learning and decision-making process. Process noise represents the uncertainty in the evolution of the hidden state (reward mean) over time. It accounts for how the true state evolves from one point in time to the next. In mathematical terms, process noise is part of the state transition equation in the Kalman Filter:

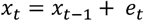

*x*_*t*_ is the state at time *t*

*e*_*t*_ is the process noise *t*, which is assumed to be drawn from a normal distribution with zeros mean and **process noise variance** *v*. Where the *e*_*t*_∼*N*(0, *v*).

The process noise captures the idea that the reward means for each arm can change from one trial to the next, even in the absence of any observations. A higher process noise variance *v* indicates a more volatile environment, where the reward means are expected to change more rapidly.

In contrast, **observation noise** represents the uncertainty in the observed rewards, given the current hidden state (reward mean). Which is assumed to be Gaussian with zero mean and a fixed variance *σ*^2^.

The observation noise captures the idea that the observed rewards are noisy and can deviate from the true reward mean due to random fluctuations or measurement errors.

A higher measurement noise variance indicates a more stochastic environment, where the observed rewards are less reliable and informative about the underlying reward means.

The Kalman Filter operates optimally when the statistical properties of the process noise and the measurement noise are accurately known.

When observation noise variance (*σ*^2^) is high relative to the process noise variance (*v*), the Kalman gain will be small, and the model will rely more on its prior beliefs and less on noisy observations. Conversely, when the observation noise variance (*v*), is high relative to the process noise variance (*σ*^2^), the Kalman gain will be large, and the model will update its beliefs more strongly based on the observed rewards.

#### Extended Kalman filter for the three-armed bandit task

The Kalman filter model can be extended to capture the effects of both volatility and stochasticity in a multi-armed bandit task (Chakroun et al., 2020; Piray and Daw, 2020). In the current study, process noise variance (□) and observation noise variance (*σ*^2^) represent volatility and stochasticity, respectively.

A traditional assumption of the Kalman filter is that the process noise variance, *v*, as well as the observation noise variance, *σ*^2^are constant.

Reward means update:

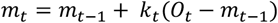

Where *m*_*t*_ is the estimated mean or value of the chosen arm at time *t* and *O*_*t*_ is the observed reward at time □.

The mean update is driven by the prediction error, which is the difference between the observed reward and the previous estimate.

Kalman gain is defined as:

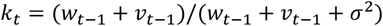

Here, *k*_*t*_ represents the Kalman gain or learning rate, which adjusts the weight given to new information based on the relative uncertainty of the prior estimate (*w*_*t*−1_) and the total noise (*v* + *σ*^2^). When the stochasticity (*σ*^2^) is high relative to the volatility (*v*), the Kalman gain (learning rate) will be small, and the model will rely more on its prior beliefs and less on the observations. Conversely, when the volatility (*v*), is high relative to the stochasticity (*σ*^2^), the Kalman gain (learning rate) will be large, and the model will update its beliefs more strongly based on the observed rewards.

Variance update equation:

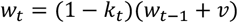

This equation updates the posterior variance (*w*_*t*_), which represents the estimate’s uncertainty after observing *O*_*t*_.

#### Volatile kalman filter for three-armed bandit task

The key difference between a standard Kalman filter and a volatile Kalman filter (VKF) is the variance of the process noise, a stochastic variable that changes with time. In other words, the VKF introduces parameters to handle the volatility in the process noise. Specifically, it allows the process noise variance *v* to vary with the observed prediction errors, reflecting changes in environmental volatility.

Kalman gain:

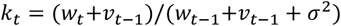

where W is a noise parameter specific to binary observations. *σ*^2^ capture the observation noise. Update for the reward means:

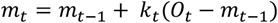

Update for posterior variance *w*_*t*_:

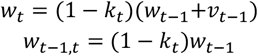

Update for volatility:

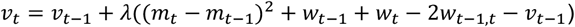

The model has three free parameters:

Where the *λ* represents the volatility learning rate, which is constrained to the unit range [0,1], determines how quickly volatility estimates update

#### Soft-max choice-probability function for (volatile) kalman filter models

Decisions were modeled using a soft-max choice-probability function in which the probability of selecting a particular bandit *i* depends on its utility.

In the first soft-max function, we only included decision weights *β*_*V*_ for expected-value. The probability P of selecting bandit *i* in trial *t* was modeled as: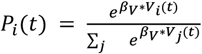

In the second response model, we only included decision weights *β*_*U*_ for outcome uncertainty to estimate the probability of choosing bandit *i*. So the probability P of selecting bandit *i* in trial *t* was modeled as:

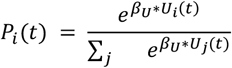

We included the third response model’s decision weights for both expected value and outcome-uncertainty. So the probability P of selecting bandit *i* in trial *t* was modeled as:

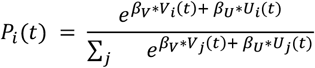

#### The definition for relative value and relative uncertainty

For each trial t, we computed the relative value (RV) and relative uncertainty (RU) of the chosen option compared to the unchosen options. Relative value was defined as the ratio of the chosen option’s estimated mean reward probability to the sum of all options’ value:

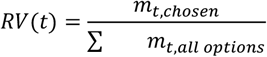

where *m*_*t*,*chosen*_ is the estimated mean of the chosen option and ∑ *m*_*t*,*all options*_ represents the sum of estimated means across all options.

Similarly, relative uncertainty was calculated as the ratio of the chosen option’s posterior variance to the sum of all posterior variances:

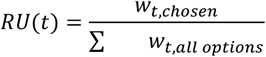

where *w*_*t*,*chosen*_ is the posterior variance of the chosen option and ∑ *w*_*t*,*all options*_ is the sum of posterior variances across all options.

### Alternative models: Rescorla-Wagner models

We also fitted the data to the classical Rescorla-Wagner model. Successful adaptation in a dynamic situation requires the appropriate feedback-based learning process where individuals integrate the feedback (reward or non-reward) into the stimulus-outcome association (Forstmann et al., 2016). The basic reinforcement learning model, the Rescorla-Wagner model can address this process well. So the first model (RW1) was defined as:

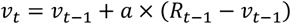

where *v*_*t*_ is the value of the option on trial *t*.

*a* represents the general learning rate from feedback.

To verify whether participants employed distinct or shared computational responses to positive and negative feedback, we built another model with two learning rates, one for positive feedback and the other for negative feedback (den Ouden et al., 2013). This model (RW2) can be defined as:

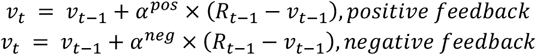

Where *v*_t_ is the value of the option on trial *t. α*^*pos*^and *α*^*neg*^ represents the learning rates from positive and negative feedback, respectively.

For these two models, *R*_*t*−1_ ∈ {0,1} represent the feedback received in response to participants’ choice on trial *t*-1. And *R*_*t*−1_ − *v*_*t*−1_ represents prediction error in trial *t*-1.

#### Soft-max choice-probability function for Rescorla-Wagner models

We used a softmax choice function to map the value into choice. The softmax function for these four models can be defined as:

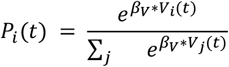

Where the *β*_*V*_ represents the inverse temperature with choice value.

### Model fitting and comparison

Hierarchical Bayesian inference (HBI) is a powerful method for model fitting and comparison in group studies (Piray et al., 2019). Unlike traditional approaches such as maximum likelihood estimation (MLE) or maximum a posteriori (MAP) estimation, which fit models to each subject independently, HBI simultaneously fits models to all subjects while constraining individual fits based on group-level statistics (i.e., empirical priors). This approach yields more robust and reliable parameter estimates, particularly when individual subject data is noisy or limited. In our study, we employed HBI to fit models to choice data. The method quantifies group-level mean parameters and their corresponding hierarchical errors. To ensure that parameter estimates remain within appropriate bounds during the fitting process, we used the sigmoid function to transform parameters bounded in the unit range or with an upper bound and the exponential function to transform parameters bounded to positive values. The initial parameters of all models were obtained using a MAP procedure, with the initial prior mean and variance for all parameters set to 0 and 6.25, respectively, based on previous research (Piray and Daw, 2020). This initial variance allows parameters to vary widely without substantial influence from the prior.

We used Bayesian model selection (26) for model comparison, specifically employing the exceedance probability (XP) to select the winning model. The XP quantifies the probability that a given model is more frequent in the population than all other models under consideration while accounting for the possibility that the observed differences in model evidence may be due to chance (see Table S1).

### Model validation

To validate our model fitting procedure, we conducted a comprehensive parameter recovery analysis that strictly follows the standard method described in the original methodology paper of VKF (Piray and Daw, 2020) (page 21/26). We generated synthetic data from 30 artificial subjects using the binary VKF combined with a softmax choice model. We employed the same Hierarchical Bayesian Inference (HBI) approach used in our empirical data analysis for parameter estimation. We repeated this procedure 20 times with different random seeds to ensure robust estimation and minimize the effects of random variation.

The recovery analysis revealed strong correlations between the true and recovered parameters, with median correlation coefficients across the 20 simulations: *r*_*λ*_ = 0.852, *r*_*v*0_ = 0.473, *r*_*σ*_^2^ = 0.919, *r*_*β*1_ = 0.918, *r*_*β*2_ = 0.782 (Figure S3).

### Control analyses for initial volatility parameter(*v*_*o*_)

While our main analyses treated the initial volatility parameter (*v*_*o*_) as a free parameter, we observed a relatively lower recovery rate compared to the other parameters. Here we demonstrate that this lower recovery rate does not compromise the model’s reliability or our main conclusions. We validate our parameter by systematically analyzing parameter recovery across different fixed values (=[1, 3, 5, 7, 9]). Using the same set of simulated data, we found that the recovery rates for the key parameters (λ, *σ*^2^, β1, β2) remained stable regardless of the fixed value. Specifically, the correlations between true and recovered parameters maintained consistent levels (λ= 0.852, *σ*^2^= 0.949, β1= 0.937, β2= 0.831) across all values, suggesting that it does not substantially interact with the recovery of other parameters.

### Exponential model analysis

In addition to linear regression analysis, we fitted exponential models to characterize the temporal dynamics of learning parameters.

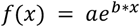

Where:

f(x) = parameter value (volatility, uncertainty, or learning rate) at trial x; a = initial amplitude coefficient (parameter value at trial 0); e = Euler’s number (≈ 2.71828); b = growth/decay rate parameter (b < 0: exponential decay (adaptive learning); b = 0: no change (flat trajectory); b > 0: exponential growth (maladaptive trajectory))

x = trial number (1 to 200)

The time constant τ = -1/b (for b < 0) represents the number of trials required for the parameter to decay to 1/e (≈37%) of its initial value. Exponential models were fitted using MATLAB’s Curve Fitting Toolbox (fit function with “exp1” model type).

See Table S2 for results.

### Control analyses

No significant correlations were detected between GCPS severity scores and uncertainty adaptation indices in TMD patients (p=0.765). Further correlation analyses revealed no significant associations between GCPS and several psychological and pain-related measures, including Apathy-Motivation Index (AMI) scores(p=0.552), Patient Health Questionnaire-9 (PHQ-9) depression severity(p=0.161), Pain Catastrophizing Scale total scores(p=0.385), and EuroQol-5D (EQ-5D) health utility indices(p=0.480). These findings suggest that, within this TMD patient sample, pain severity and disability, as measured by GCPS, operate independently from uncertainty processing mechanisms and related psychological factors. Correlation analyses were performed using Pearson’s correlation coefficient (Figure S4).

## Data Availability

Anonymized behavioral, model-derived data, and analysis code are available upon request.

## Data, Materials, and Software Availability

Anonymized behavioral, model-derived data, and analysis code are available upon request.

## Acknowledgments

This work was supported by NIMH under award #R21MH127607, NIDA under award #K23DA050909, and the University of Minnesota’s MnDRIVE (Minnesota’s Discovery, Research and Innovation Economy) initiative. We thank the participants, staff, and providers at the University of Minnesota TMD, Orofacial Pain and Dental Sleep Medicine Clinic for their contributions.

