## Supplementary Information for "Impaired Adaptive Learning in Chronic Pain Contributes to Apathy"

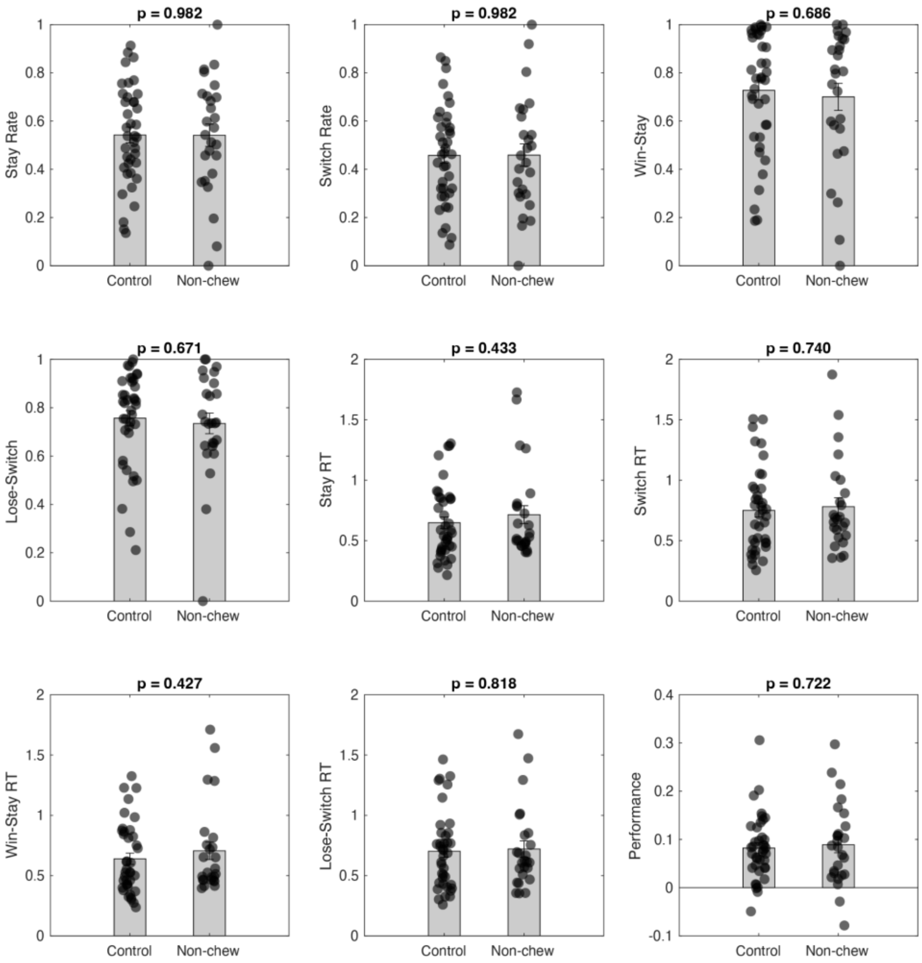


**Figure S1.** Model-free behavioral metrics show equivalent task performance between TMD patients and healthy controls.

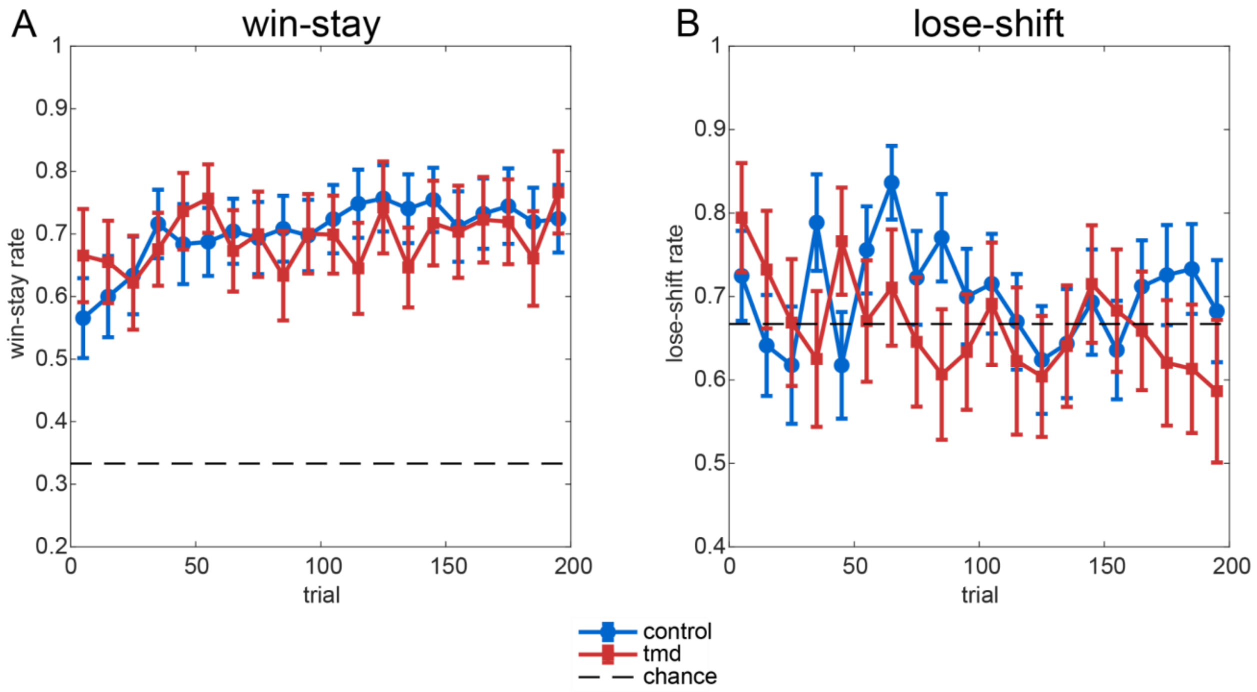


**Figure S2. Temporal dynamics of behavioral metrics across learning.**

Win-stay rate (left), lose-shift rate (middle), and performance (right) shown in 10-trial bins across the 200-trial task. Blue circles represent controls (n=39), red squares represent TMD participants (n=26). Error bars indicate SEM. Dashed horizontal lines indicate chance levels (0.333 for win-stay, 0.667 for lose-shift). Both groups maintain stable behavioral performance throughout the task without systematic group differences.

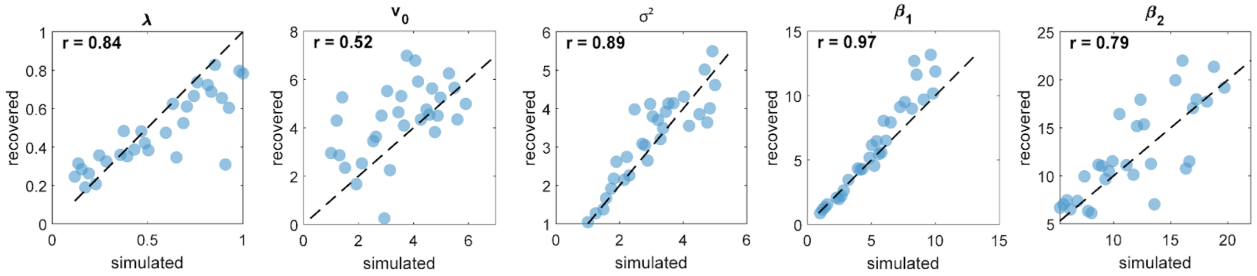


**Figure S3. Parameter recovery analysis.**

The scatter plots show the relationship between simulated (true) and recovered parameters for the binary VKF model with softmax choice rule. Each point represents one artificial subject (n=30). The dashed lines indicate perfect recovery (y=x). Correlation coefficients (r) are shown for each parameter. Parameters shown are step size (λ), initial volatility parameter($v_0$ ), observational noise ($\sigma^2$), and choice sensitivity parameters ($\beta_1$ , $\beta_2$ ). Note: Parameters were estimated using Hierarchical Bayesian Inference (HBI). Correlations indicate strong parameter recovery for most parameters, with moderate recovery for initial uncertainty ($v_0$ ).

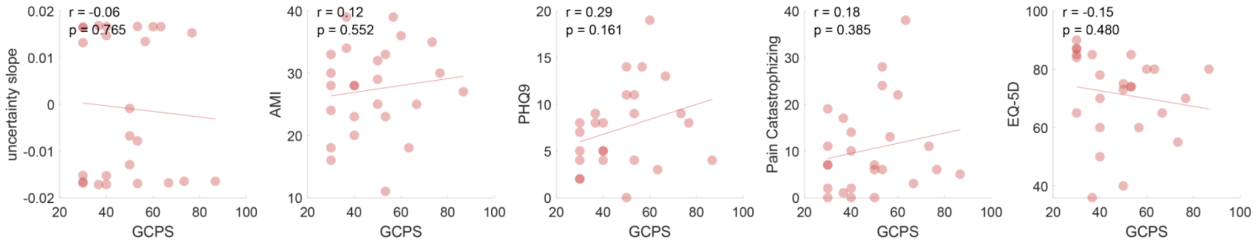


**Figure S4. Correlation analyses between the Graded Chronic Pain Scale (GCPS) and uncertainty adaptation, and other clinical measures within the TMD patient cohort.**

No significant correlations were detected between GCPS severity scores and uncertainty adaptation indices in TMD patients. Further correlation analyses revealed no significant associations between GCPS and several psychological and pain-related measures, including Apathy-Motivation Index (AMI) scores, Patient Health Questionnaire-9 (PHQ-9) depression severity, Pain Catastrophizing Scale total scores, and EuroQol-5D (EQ-5D) health utility indices. These findings suggest that, within this TMD patient sample, pain severity and disability, as measured by GCPS, operate independently from uncertainty processing mechanisms and related psychological factors. Correlation analyses were performed using Pearson’s correlation coefficient.

***Table S1. Model performance***

| All models | XP (exceedance probabilities) (TMD patients) | XP (exceedance probabilities) (HC) |
|---|---|---|
| RW1 | 0 | 0 |
| RW2 | 0 | 0.0794 |
| KF | 0 | 0 |
| VKF-RV | 0 | 0 |
| **VKF-RVRU** | **1.000** | **0.9206** |

**Notes:** Model comparison results using exceedance probabilities (XP). Higher exceedance probabilities indicate better model fit. RW1 = standard Rescorla-Wagner model with single learning rate; RW2 = Rescorla-Wagner model with separate learning rates for reward and no-reward; KF = standard Kalman filter; VKF-RV = volatile Kalman filter with relative value; VKF-RVRU = volatile Kalman filter incorporating both relative value and relative uncertainty. The VKF-RVRU model showed the highest XP, indicating that it best explains participants' choice behavior.

***Table S2. Results from the exponential model analysis***

| **parameter** | **group** | **decay (b<-0.001)** | **growth (b>0.001)** | **flat** | **mean b ± SEM** | **mean $R^2$** |
|---|---|---|---|---|---|---|
| volatility | HC | 32 (82.1%) | 4 (10.3%) | 3 (7.7%) | -0.0101 ± 0.0047 | 0.814 |
| | TMD | 11 (42.3%) | 10 (38.5%) | 5 (19.2%) | -0.0022 ± 0.0018 | 0.786 |
| | HC vs. TMD | *p = 0.001** | | | *p = 0.189* | |
| uncertainty | HC | 18 (46.2%) | 1 (2.6%) | 20 (51.3%) | -0.0009 ± 0.0001 | 0.808 |
| | TMD | 7 (26.9%) | 8 (30.8%) | 11 (42.3%) | +0.0001 ± 0.0003 | 0.773 |
| | HC vs. TMD | *p = 0.193* | | | *p = 0.002* | |
| learning rate | HC | 27 (69.2%) | 2 (5.1%) | 10 (25.6%) | -0.0018 ± 0.0003 | 0.779 |
| | TMD | 9 (34.6%) | 8 (30.8%) | 9 (34.6%) | -0.0006 ± 0.0006 | 0.760 |
| | HC vs. TMD | *p = 0.010* | | | *p = 0.039* | |

Note. *p-values for decay proportions from Fisher's exact test; p-values for b parameters from two-sample t-tests
